# App-based Epidemic Game to Model Belief-Behavior Mapping and Cost Incentives in Voluntary Quarantine: A Randomized Controlled Trial

**DOI:** 10.64898/2026.01.10.26343836

**Authors:** Andrés Colubri, Andonaq Grozdani, Mansi Khandpekar, Charvi Bansal, Yousif Graytee, Omar Al-Mohammedi, Ahmed Ayden Al-Shabandar, Wid Yasir Shabeeb, Yaqoob Ghassan, Hayder Swayedi, Chris T. Bauch, John Drury, Jasmina Panovska-Griffiths, Dmitri Williams, Dale King

## Abstract

**Background:** Understanding protective behavior during infectious disease outbreaks is critical for public health policy. App-based experimental epidemic games (epigames) offer a novel empirical approach, but their external validity and feasibility as rigorous randomized-trial platforms require evaluation. We examined whether real-life health beliefs transferred into an epigame, whether opportunity cost influenced voluntary quarantine, and whether the design could support subsequent behavioral experiments.

**Methods:** We conducted a preregistered 15-day randomized controlled trial using the Epigames smartphone app at the American University of Iraq – Baghdad, with a total of 539 registered participants. The app sampled Bluetooth proximity at sub-minute resolution, simulated respiratory-pathogen transmission through the contact network, and elicited daily quarantine choices. Participants were randomized to a low-cost barrier, receiving 5 points for quarantine and 10 for remaining active, or a high-cost barrier, receiving 5 and 17 points, respectively; a willingness-to-accept pilot informed the contrast. Real-life (S1) and in-game (S2) surveys measured susceptibility, severity, quarantine self-efficacy, and perceived benefits. We assessed belief correspondence with Holm-adjusted Spearman correlations and prospective cumulative-logit models and analyzed choices using participant-day binomial generalized estimating equations (GEE) and participant-level quasi-binomial models.

**Results:** The trial integrated app-based randomization, longitudinal contact measurement, repeated choices, and timed surveys in a naturalistic campus setting. Participants contributed 1,611 participant-days and 1,615 recorded choices. Among 257 participants with matched surveys, all four corresponding S1–S2 beliefs were positively associated (Spearman’s ρ=0.126–0.353; all Holm-adjusted p<0.05), and prospective ordinal models confirmed matched-domain correspondence. Quarantine rates were 9.6% under the low barrier and 8.9% under the high barrier. The high barrier did not detectably alter quarantine overall (GEE OR 0.90, 95% CI 0.58–1.40, p=0.650; quasi-binomial OR 0.93, 95% CI 0.58–1.48, p=0.753), and joint belief-by-group tests were null. Two non-confirmatory patterns warrant prospective investigation: baseline severity showed a small positive association with quarantine rate (ρ=0.132, raw p=0.0155, Holm p=0.0619), and the estimated high-barrier effect for “skeptical” participant (those with responses at the floor of all S2 belief scales) was large but specification-sensitive (GEE OR 0.060, p=0.039; quasi-binomial OR 0.058, p=0.056). Women were more likely than men ever to quarantine (arm-adjusted OR 2.76, p=0.0002), but there was no gender-by-barrier interaction.

**Conclusion:** This study demonstrates the feasibility of a preregistered epigame RCT combining randomized incentives, passively measured contacts, repeated protective choices, and prospectively measured beliefs. The strongest evidence concerned real-life to in-game belief correspondence; the severity and low-belief barrier patterns are targets for adequately powered, prospectively specified replication. Technical limitations, uneven prompt response, and gameplay that may not have sustained engagement could have attenuated behavioral contrasts. Epigames provide a promising foundation for network-aware behavioral epidemiology. Protocol refinement and multisite replication are needed before inferring real-world behavioral validity.

**Study pre-registration:** https://osf.io/qev6n/

## 1. Introduction

Human behavior is a decisive factor in the dynamics of infectious disease transmission [1–6]. However, quantifying the causal impact of behavioral changes, such as voluntary quarantine, on pathogen spread remains a fundamental challenge in epidemiology. Observational data from real-world outbreaks are often confounded by simultaneous interventions and varying compliance, while mathematical models frequently rely on assumed rather than empirical parameters for human decision-making. True experiments are impossible for clear ethical reasons. Consequently, there is an urgent need for experimental frameworks that can isolate and quantify the drivers of protective behaviors under controlled yet realistic conditions.

Experimental epidemic games, or “epigames,” conducted in naturalistic settings, are inspired by behavioral economics and game theory, and offer a promising avenue to fill this gap [7–9]. By placing participants in social dilemmas within a realistic framing where personal decisions impact both individual and collective outcomes, researchers can simulate the incentives inherent in disease prevention and mitigation [10–14]. While controlled behavioral experiments, often relying on web-based surveys and incentivized choice games [15–17] are designed to satisfy internal validity, they may lack external validity, specifically, by not being able to replicate the complex social contexts and realistic disease exposure patterns of daily life given their simplified framing and the fundamental issue of different real costs such as sickness and death. Conversely, early digital epidemiology projects like the “Corrupted Blood” incident in World of Warcraft [18] or the FluPhone project [19] demonstrated that highly immersive or sensor-based environments could generate realistic contact networks and rich behavioral data, and provide the basis for the epigames approach developed here. However, critical questions remains: *do the decisions players make in these gamified proxies “map” over to their real-life health beliefs, or are they merely artifacts of the game mechanics?* [20] *Can app-based gamified experiments reach a level of immersion and incentive that is verifiably parallel to their real-life counterparts?*

Despite the potential of the experimental games in general to provide insights into real-world behavior and cross-cultural differences often not captured by traditional ethnographic or survey methods [21], they have also been criticized for lacking external validity [22–26]. Anthropologists have responded to these critiques by testing framing effects [25, 27] and minimalism in experimental designs [23]. Although debates continue about the universal applicability of experimental game results, these instruments can provide meaningful insights into human responses under well-defined experimental conditions. Much like accepted disaster preparedness exercises, including the “Great ShakeOut” earthquake drills [28] and the Humanitarian Response Intensive Course [29], experimental games serve as performative “stress tests” that reveal behavioral patterns and systemic vulnerabilities within a safe, controlled environment.

It’s important to distinguish two dimensions of external validity: generalizability, whether observed behaviors hold across populations and settings, and parallelism, the extent to which in-game mechanisms (context parallelism, or ecological validity) and behaviors (indicator parallelism) reflect real-world counterparts [30, 31]. Given the question of whether epigames can be used to study preventive health behaviors, we are interested in maximizing context and indicator parallelism as critical measures of external validity. More specifically, indicator parallelism is vital for establishing epigames as a rigorous scientific tool. If epigames are to serve as “laboratories” for public health policy, we must demonstrate that a participant’s in-game choice to quarantine is driven by the same psychological factors (e.g., perceived susceptibility, severity, and efficacy) that drive real-world compliance. This validation is particularly crucial for understanding high-stakes interventions like quarantine, where individuals must weigh the collective public health benefit against immediate personal and economic costs.

To address these challenges, we developed the *Epigames* smartphone app [32] based on the *Operation Outbreak* (OO) app that introduced the use of Bluetooth proximity sensing in experiential learning simulations of infectious disease spread [33]. Building on this foundation, the *Epigames* app allows us to conduct large-scale, randomized trial studies within a high-resolution, large-scale epidemic simulation. Unlike previous retrospective or exploratory analyses of contact networks and human behavior [34, 35], this study was explicitly designed to investigate the relationship between pre-existing real-life health beliefs and in-game protective choices, specifically examining how economic barriers (opportunity costs) moderate this relationship in the case of voluntary quarantine.

We designed the study as a randomized controlled trial (RCT). The app recorded real-world contact networks via Bluetooth to simulate the spread of a “digital” respiratory virus and represented health status via an animated avatar. However, “quarantine” was operationalized as a digital decision rather than physical isolation (which would have been difficult to implement): participants could choose to suspend their Bluetooth contacts via an in-app button, thereby preventing infection. To simulate real-world economic and social costs, choosing to quarantine incurred a point penalty compared to remaining active. We acknowledge that this digital abstraction is a much-simplified proxy for the complex burden of physical isolation that limits context parallelism of our study, but it allows for the precise experimental manipulation of the economic barrier. Participants were randomized into two arms: a Low Barrier Group (G1) facing a nominal penalty, and a High Barrier Group (G2) facing a significant penalty. Participants completed two surveys measuring health belief constructs, first regarding real-life respiratory diseases (S1), and subsequently regarding the game context (S2), to assess the external validity of their in-game beliefs and behaviors. Consistent with our pre-registration on the Open Science Framework (OSF) website (https://osf.io/qev6n/) [36], we tested three hypotheses: (H1) Main Effect: G1 will exhibit higher quarantine rates than G2; (H2) Moderation by Cost: The correlation between personal beliefs and behavior will be stronger in G1, as high costs in G2 overpower personal motivations; and (H3) Interaction by Efficacy: The lower quarantine rates due to high barrier in G2 will be most pronounced among participants with low self-efficacy. In addition to these hypotheses, surveys S1 and S2 allowed us to investigate several research questions. (RQ1) Real-Life to Game mapping: is there a significant correlation between S1 and S2 variables? (RQ2) Demographics and Quarantine: are demographic variables collected in S1, such as gender, associated with the quarantine decision during the game? Finally, the measurement of the participants’ contact network via Bluetooth enabled an additional research question. (RQ3) Network Structure: what topology best describes the contact network and whether the properties of its nodes exhibit assortative mixing.

We adopted the Health Belief Model (HBM) [37, 38] as an initial, although not definitive, theoretical framework for this investigation. A reason for doing so is HBM’s distinction of being one of the most utilized models for predicting health behaviors [39], which makes it a well-known and understood model in the field of health behavioral research. HBM posits that an individual’s decision to adopt a protective action such as voluntary quarantine is determined by the interplay of specific cognitive perceptions: perceived susceptibility to the illness, perceived severity of its consequences, perceived benefits of the action, and perceived barriers to performing it. In the context of infectious disease, the model suggests that compliance is a calculated trade-off where the threat of infection is weighed against the tangible costs of safety measures. In our study, we specifically operationalize “perceived barriers” through the game’s economic opportunity costs, allowing us to experimentally test how high-cost barriers suppress protective behaviors even when perceived risk is held constant. By constructing a conceptual HBM using the data from the AUIB epigame, we aimed at demonstrating the use of the novel epigame approach to quantify how economic costs act as a barrier to protective behaviors [40, 41], providing empirical evidence to inform pandemic preparedness strategies.

## 2. Methods

### 2.1 Study Design and Participants

We recruited 567 participants from the American University of Iraq – Baghdad (AUIB) in the Middle East. A target sample size of N=500 was determined as a conveniency sample based on earlier pilots and total school enrollment (∼2,500 students). Eligibility criteria included being a student, faculty, or staff member at AUIB, being at least 18 years of age, and possessing an iOS or Android smartphone capable of running the *Epigames* app. The study was a 15-day, two-arm, parallel-group, single-blind randomized controlled trial. Randomization was performed by the app using a coin-flip algorithm upon registration, without researcher involvement.

Of the total 567 registered participants, 545 had a recoverable group assignment (246 G1 and 299 G2), including nine assignments reconstructed from auxiliary study records; 22 records without a recoverable assignment due to randomization error in the app were excluded. Six participants withdrew, leaving 539 retained records (244 G1 and 295 G2). A further 136 participants made no recorded quarantine decision. The primary behavioral analysis therefore included 403 participants who made at least one decision (188 G1 and 215 G2). Analysis populations varied according to survey completion and are reported for each model. Among participants with S1 gender data used in binary analyses, 158 identified as men and 169 as women; participants reporting another category or without a usable gender response were excluded only from analyses requiring the binary variable. Refer to **Figure 1A** for the study’s CONSORT flow diagram.

**Figure 1:**
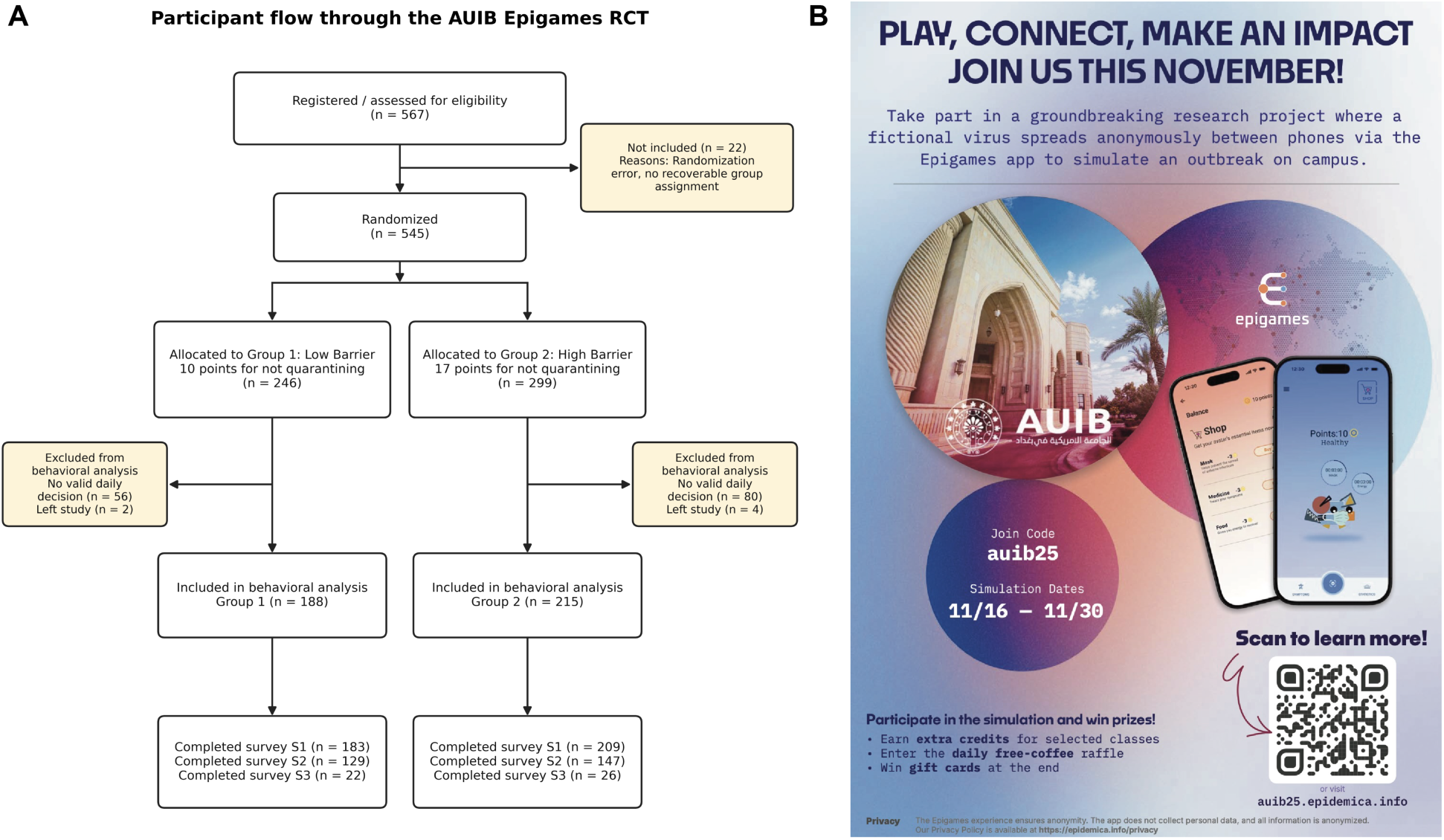
Study implementation and participant recruitment. The panel on the left (A) contains the CONSORT flow diagram. Of 567 registered records, 545 had a recoverable group assignment, six participants withdrew, and 403 participants made at least one recorded quarantine decision and entered the behavioral analysis. The panel on the right (B) shows the flier used to promote the epigame on the AUIB campus.

A month-long recruitment campaign comprising an information desk with physical fliers, an informational website and emails, class presentations, and a Telegram chat group, was conducted on campus by a local team of student volunteers (**Figure 1B**). Participants provided informed consent after installing the app to join the epigame and were incentivized via daily raffles for local store vouchers to maintain engagement. Points collected through the app were used in a lottery for gift cards, with the number of tickets proportional to the participants’ final point score. The study protocol was reviewed and approved by UMass Chan IRB (ID 2465) and received clearance by AUIB IRB.

### 2.2 The Epigames app

The study utilized the *Epigames* app, which simulates a “digital pathogen” transmitting between nearby users of the app. This app was built upon the existing OO app [15] for experiential learning simulations on infectious disease [16]. The *Epigames* app uses the same Bluetooth proximity sensing technology as OO, based on the open-source Herald project [17], to sample high-resolution contact networks between participants, while adding additional infrastructure for gamified behavioral studies, including the assignment of participants to randomized groups, pre-, mid-, and post-game surveys, and gamified interventions such as quarantine and contact tracing (whose parameters may depend on group randomization).

For the epigame at AUIB, the digital pathogen in the app was parametrized to mirror the properties of Omicron variant of SARS-CoV-2 with an R0 of ∼7 [42]. Duration of incubation period, serial interval, and recovery rate were calculated to fit approximately three consecutive waves of disease spread in the two weeks of the epigame. Similar parameters were used in an earlier pilot outbreak simulation with the OO app at an international college campus [35]. This pilot yielded highly realistic transmission trees with super-spreader statistics closely matching those of real-life COVID-19 transmission trees [43]. The epigame was seeded by the app randomly choosing several index cases among the participants.

### 2.3 Belief Measures

The app allows multiple-choice surveys to be administered to participants at any point during the study. For the epigame at AUIB, we constructed three surveys to evaluate relationships between prior real-life beliefs and in-game behavior (pre-registered hypotheses H1–H3), correspondence between real-life and in-game beliefs, demographic associations with in-game behavior (research questions RQ1 and RQ2), and participants’ post-game self-assessments:

- **S1 (Real-Life Beliefs):** Baseline survey measuring perceived susceptibility (Q1) and severity (Q2) about respiratory viruses, and self-efficacy (Q3) and benefits (Q4) regarding voluntary quarantine, all in real-world settings, and basic demographics (gender, school affiliation). It was administered in the first day of the game, shortly after informed consent and before any transmission has occurred. It was done this way since no data collection was possible before consent, but early enough to minimize potential biases resulting from the study itself.
- **S2 (In-Game Beliefs):** Survey measuring the same constructs as in S1 (susceptibility, severity, self-efficacy, and benefits) but now reframed in the context of the epigame (*“If your avatar gets infected with the virtual pathogen…”*) to provide the necessary data for the real-life-to-game mapping calculations. It was administered at the end of the third day of the epigame, when index cases were already being assigned, but symptoms were still to be shown through the *Epigames* app. In this way, participants had time to familiarize themselves with the app so they could provide meaningful responses to this survey without being influenced by the simulated transmission or disease events.
- **S3 (Exit Survey):** Administered on the final day, this survey assessed the epigame’s external validity and user experience. Participants evaluated the realism of disease transmission and their own quarantine decisions, reported their anxiety regarding virtual infection, and provided usability feedback and additional demographic data.

These surveys are provided in the supplementary materials, and all used a Likert scale between 1 and 6, except for demographic questions (gender, school affiliation), to avoid a neutral midpoint and therefore requiring respondents to give a non-neutral view. They were also available in Arabic if the participants had their phone’s user interface set to that language as default.

### 2.4 User Interface and Gamified Quarantine Choice

Participants joined the epigame by first installing the *Epigames* app, available free of charge from the Apple and Google app stores, then entering a code that uniquely identified the AUIB epigame, granting access to Bluetooth and other services required by the app, and finally providing informed consent digitally (**Figure 2**). Throughout the duration of the epigame, participants were updated about their simulated health status through an animated avatar on the main screen on the app. Each participant was randomly assigned a distinct character for their avatar from a set of five different, pre-determined characters. The whimsical design of these characters was inspired by the concept of a “digital pet” [44, 45] to elicit user attachment with their avatar, increase engagement during the game, and potentially strengthen the mapping between real-life and in-game behaviors [46]. Feedback from earlier pilots suggested that users found these avatars pleasant and some requested making them more interactive and customizable, which opens possible paths for future improvements in the app’s user experience. Beyond this aspect, the *Epigames* app offered other interface features, including a screen displaying nearby contacts and their respective health and quarantine states. The surveys were administered through the app via push notifications. Upon tapping on these notifications, participants had to complete the associated survey to continue using the app.

**Figure 2:**
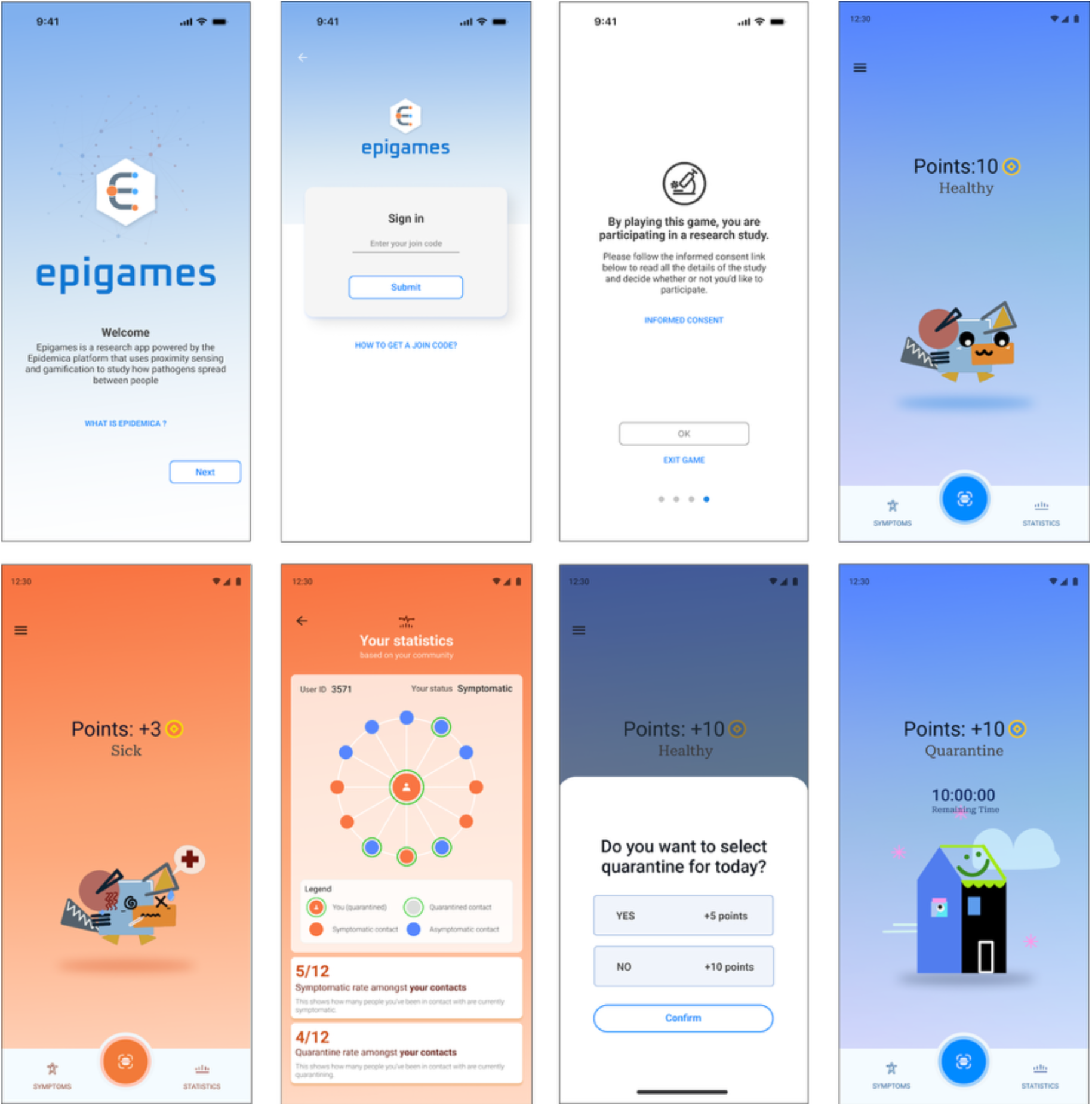
Epigames app. The images in the top row depict, from left to right, the welcome, sign in, informed consent, and home screens. The avatar in the home screen conveys the health and quarantine status of the user. In the bottom row, from left to right, the home screen displaying the avatar in the sick state, the statistic screen with information of neighboring players and their respective statuses, the popup with daily quarantine choice, and home screen with avatar in quarantined state.

A key feature in this epigame was the quarantine choice. This was implemented with a popup in the app that was presented daily to all participants between 8:00 AM and 1:00 PM. To model the burden of real-world quarantine without requiring physical isolation, we operationalized “cost” through a differential point-based incentive structure:

1. **Quarantine:** Isolate the avatar to prevent infection due to interactions in the real world (by suspending the app’s Bluetooth proximity sensing). The participant receives the lowest number of points by making this choice (Low Reward). This point penalty serves as a digital proxy for the real-life costs of quarantining (e.g., lost income, social isolation), operationalizing the "barrier" to safety.
2. **Do Not Quarantine:** Risk infection of the avatar by allowing interactions in the real-world as measured via Bluetooth, but the participant receives a larger number of points (High Reward), representing the immediate utility and benefit of maintaining normal social and economic activity.

Infection resulted in “illness” (during which point rewards were reduced to a minimum irrespective of quarantine choice), clearly represented by changes in app’s UI color and avatar appearance. Illness could lead to “death” of the avatar and loss of all accumulated points. Total points for each participant were displayed on a public leaderboard in the app that updated daily as participants acquired points by making their quarantine choices or lost points due to illness.

We expressly designed this mechanic to abstract the multifaceted burden of quarantine into a quantifiable economic decision. While we recognize that a reduction in app points cannot replicate the physical, psychological, and social costs of actual isolation, thereby limiting the context parallelism of the study, this abstraction allowed us to experimentally manipulate the cost variable across treatment groups (G1 vs. G2) to measure behavioral elasticity.

### 2.5 Outcome measures

The primary outcome was the daily quarantine decision (binary), summarized per participant as the number of quarantine decisions and the quarantine rate across all recorded decisions during the 15-day study. The behavioral cohort comprised participants with at least one recorded choice. Four participant-days contained both a quarantine and a non-quarantine event; the participant-level analysis retained both recorded events in the denominator, whereas the participant-day construction classified a day as quarantine if any quarantine event was present.

Secondary outcomes included correspondence between matched real-life (S1) and in-game (S2) beliefs, modification of the randomized barrier effect by those beliefs, gender differences in quarantine adoption and propensity, and descriptive properties of the contact network. Daily contacts and simulated infections were summarized descriptively, and final point scores determined lottery entries after the epigame.

### 2.6 Determining the Economic Barriers to Quarantine

A “Willingness to Accept” (WTA) [47] pilot survey (N=50) was conducted prior to the main experiment. We shared an anonymous Google form via a Telegram chat regularly used by AUIB students, which presented with the following scenario and questions:

- Scenario: “In the upcoming epigame, you will make a daily choice about your in-game avatar:

- Choice A: ‘Quarantine’ your avatar. You get 5 points, guaranteed.
- Choice B: ‘Do Not Quarantine’ your avatar. This is the risky choice. You get more points, but only if you avoid your avatar from getting virtually ‘infected’. If your avatar gets infected, you risk losing all your points.”
- Questions:

- “If Choice B gave you 10 points (a 5-point gap), which would you choose? (A or B)”
- “If Choice B gave you 15 points (a 10-point gap), which would you choose? (A or B)”
- “If Choice B gave you 20 points (a 15-point gap), which would you choose? (A or B)”
- “If Choice B gave you 25 points (a 20-point gap), which would you choose? (A or B)”
- “If Choice B gave you 30 points (a 25-point gap), which would you choose? (A or B)”

Analysis of the responses was conducted to find the median acceptance threshold, defined as the point gap at which approximately 50% of the respondents “flip” from Choice A to Choice B. This identified the point of maximum behavioral tension at a gap of 12 points (see **Suppl. Figure 1**). Consequently, participants were randomized into:

- **Group 1 (Low Barrier):** In this group, participant who quarantined received 5 points (Low Reward), those who did not received 10 points (High Reward), therefore the economic barrier to quarantine was 5 points (nominal/low).
- **Group 2 (High Barrier):** In this group, participant who quarantined received 5 points (Low Reward) as before, but those who did not, received 17 points instead (High Reward), therefore the economic barrier to quarantine was 12 points (high).

### 2.7 Statistical Analysis

Analyses followed the pre-registered hypotheses; with variance estimation revised after diagnostic analyses showed extra-binomial variation and within-participant association in repeated daily choices. All tests were two-sided, estimates are reported with 95% confidence intervals (CIs), and statistical significance was assessed at α=0.05.

Correspondence between real-life and in-game beliefs (RQ1) was assessed using Spearman rank correlations between the four prespecified matched S1–S2 item pairs: susceptibility, severity, self-efficacy, and perceived benefits. Family-wise error across these four correlations was controlled using the Holm procedure. Because responses were ordinal, prospective cumulative-logit models were also fitted for each S2 item, first using its matched S1 item and randomized group and then using all four S1 items and group. Marginal associations between individual belief items and participant quarantine rate were treated as exploratory and summarized with Spearman correlations.

Population-averaged generalized estimating equation (GEE) models were fitted to participant-day binary decisions using a binomial family, logit link, participant clustering, exchangeable working correlation, and robust standard errors [48]. In parallel, participant-level quarantine proportions were analyzed using binomial generalized linear models with logit link, weights equal to the number of recorded decisions, and Pearson quasi-likelihood scaling (scale=’X2’) to accommodate extra-binomial heterogeneity [49]. The GEE represented the disaggregated repeated-decision analysis, whereas the quasi-binomial model represented overall participant quarantine propensity while retaining each participant’s denominator. Agreement between these complementary analyses was used to assess robustness. The unscaled Poisson and binomial models in the original analysis plan were not used for final inference because their variance assumptions underestimated the observed variation.

The pre-registered H1 overall barrier effect was estimated from models containing randomized group alone. Models for H2 and H3 included the four baseline S1 beliefs, binary gender, randomized group, and all belief-by-group interactions. The four interactions were tested jointly for H2, and the self-efficacy-by-group term tested H3. Corresponding models using S2 beliefs were treated as secondary in-game extensions. Belief items were mean-centered for the fitted models. Separate linear contrasts estimated the randomized group effect for a participant scoring 1 on all four belief items (the “skeptic” profile), standardized over the gender distribution of the relevant analysis sample. These floor-profile contrasts did not replace the pre-registered overall H1 test. Complete-case S1 and S2 interaction models included 327 and 250 participants, respectively.

Gender was coded as 0 for men and 1 for women. The gender main effect in multivariable belief models, the gender-by-group interaction, and group-specific estimates were used to distinguish overall gender differences from differential barrier effects. Because repeated-choice and participant-level specifications gave different levels of support for the gender main effect, adoption of quarantine at least once was also examined using participant-level logistic regression. Gender analyses were interpreted with attention to this specification dependence.

As sensitivity analyses, the overall group effect was estimated using fractional logistic regression with HC1 heteroskedasticity-robust standard errors and ordinary least squares regression of participant quarantine rate with HC3 standard errors. Pearson dispersion, the association between consecutive choices, and the exchangeable GEE working correlation were used as specification diagnostics.

The aggregate contact network pooled Bluetooth proximity events across the study. Nodes represented participants and weighted edges represented cumulative contact. Network topology and assortativity were summarized descriptively [50]. Assortativity analyses were considered exploratory because multiple attributes were examined and the resampling procedure did not preserve all forms of network dependence; no confirmatory claims were based on their nominal p-values.

All statistical analyses and plots were performed with Python 3.13.5 and the following packages: numpy 2.3.0, pandas 2.3.0, pyarrow 20.0.0, pygraphviz 1.14, matplotlib 3.10.3, networkx 3.5, scikit-learn 1.7.0, scipy 1.15.2, statsmodels 0.14.4, and seaborn 0.13.2. The code repository is provided in the supplementary materials.

## 3. Results

### 3.1 Participation and Daily Game Activity

The behavioral cohort included 403 participants (188 G1; 215 G2). Complete S1 beliefs were available for 336 participants, complete S2 beliefs for 263, matched S1–S2 beliefs for 257, and complete S3 responses for 48. Participants contributed 1,611 participant-days and 1,615 recorded choices. Overall, 149 choices were quarantine and 1,466 had remained active; 26.3% of participants quarantined at least once.

Bluetooth activity followed the campus week, with lower activity on Thursdays and the Friday–Saturday weekend. Simulated infections generally followed peaks in recorded contacts. Choice response volume also varied markedly across days, and remaining active was more common than quarantine throughout the study. These trends are shown in **Figure 3**.

**Figure 3:**
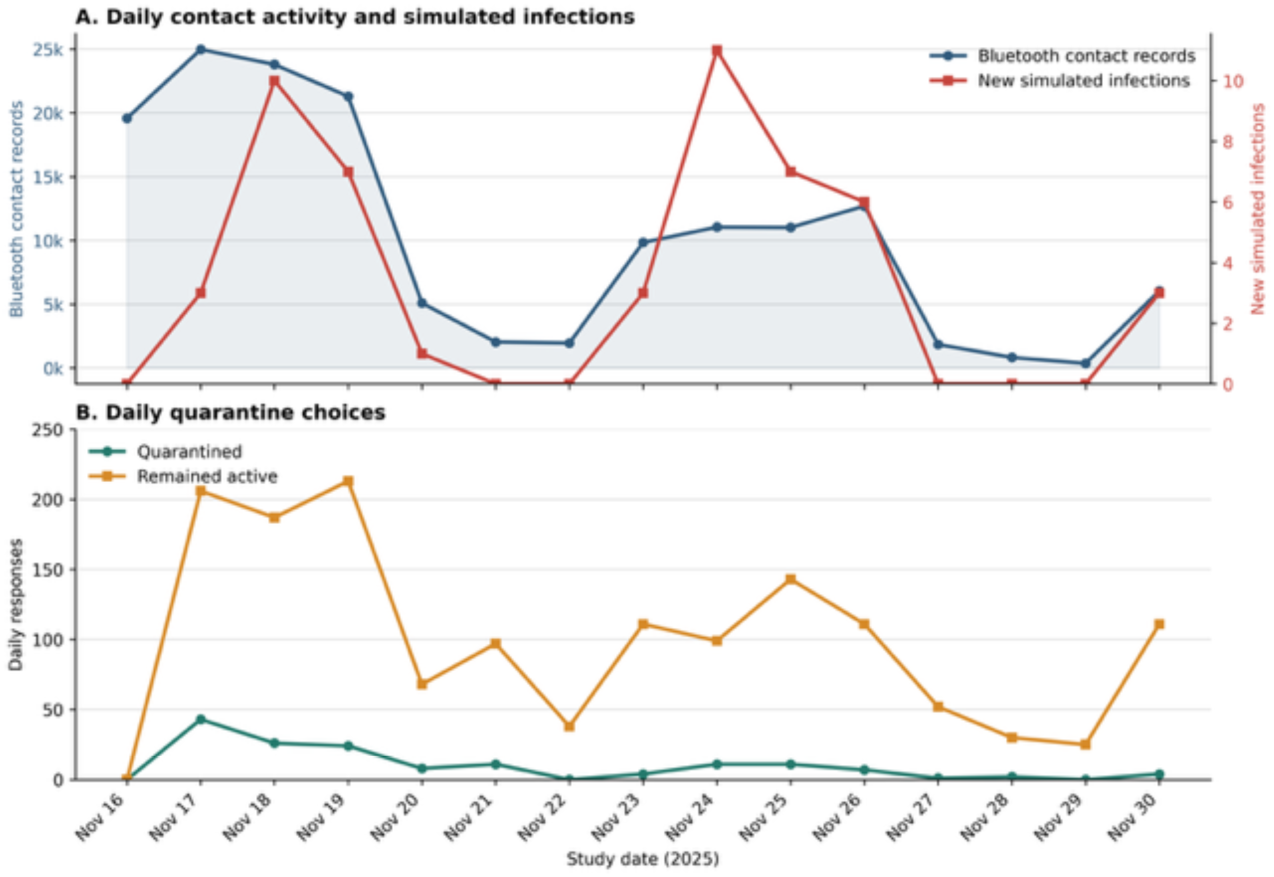
Daily game activity and behavioral choices. (A) Bluetooth contact records and new simulated infections by study day, shown on separate vertical scales. (B) Numbers of daily quarantine and remain-active choices. Counts reflect recorded responses and therefore vary with daily participation.

### 3.2 Survey Response Patterns

Complete responses to Questions 1–4 were available for 336 participants for S1, 263 for S2, and 48 for S3. The distributions differed by construct and survey framing (**Figure 4**). Because S3 represented a small, self-selected subset, its distributions are descriptive and should not be treated as population estimates.

**Figure 4:**
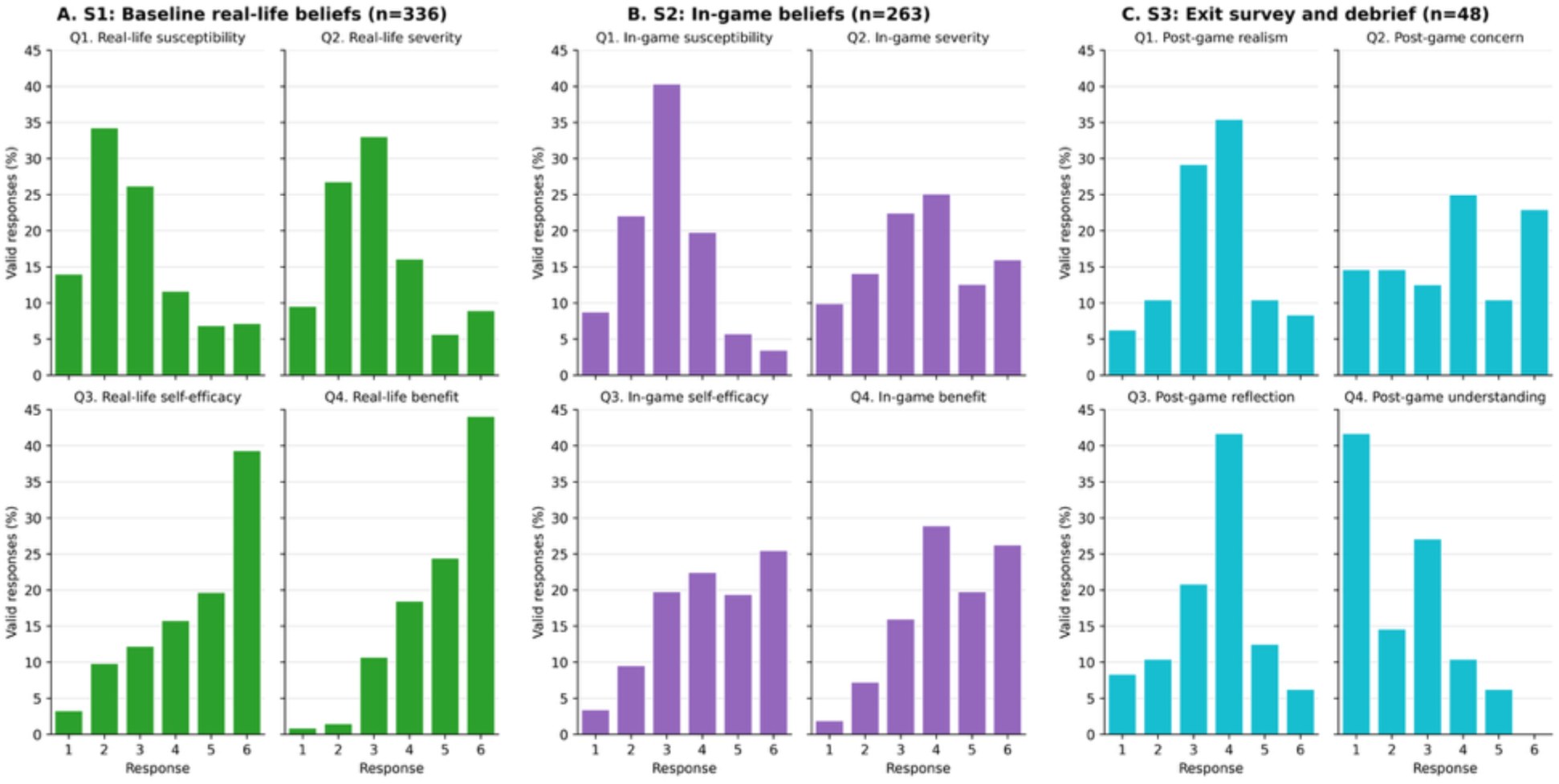
Survey response distributions. Percentages of valid responses to Questions 1–4 from (A) baseline real-life beliefs, (B) in-game beliefs, and (C) the exit survey. S3-Q4 is difficulty understanding information, so higher values indicate greater difficulty.

### 3.3 Transfer of Real-Life Beliefs into the Game

All four matched real-life and in-game belief measures were positively associated among the 257 participants with complete S1–S2 data (**Figure 5**). Spearman correlations were ρ=0.204 for susceptibility, ρ=0.126 for severity, ρ=0.267 for self-efficacy, and ρ=0.353 for perceived benefits. Holm-adjusted p-values were 0.0020, 0.0431, <0.001, and <0.001, respectively.

**Figure 5:**
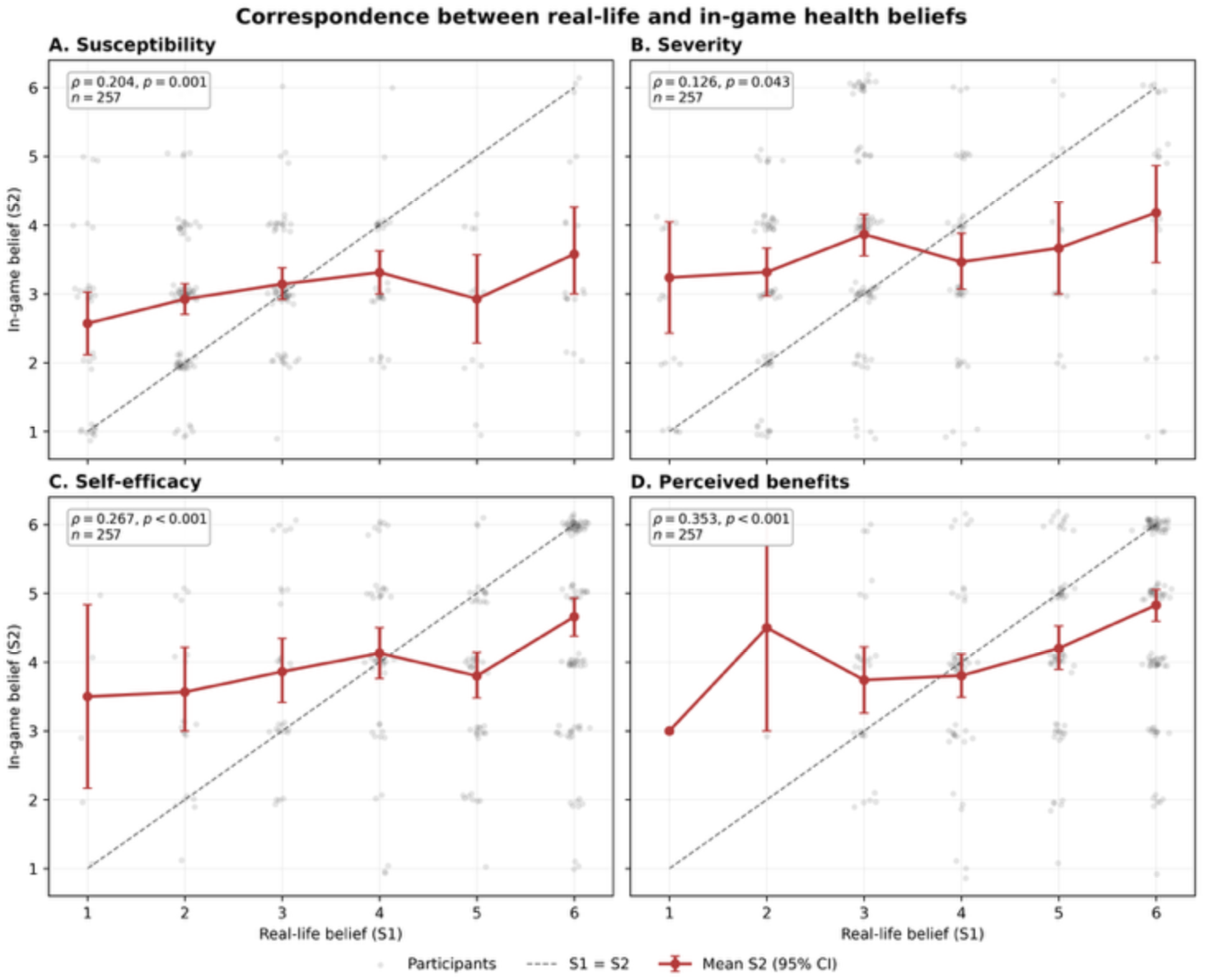
S1 Correspondence between matched real-life and in-game beliefs. Points show paired responses; summaries show mean S2 response at each S1 level with bootstrap CIs. The diagonal denotes identical responses. Correlations are Spearman rank estimates.

Prospective cumulative-logit models corroborated these associations. Per one-point increase in the corresponding S1 belief, the cumulative odds ratios for a higher S2 response were 1.32 (95% CI 1.11–1.57) for susceptibility, 1.21 (95% CI 1.02–1.44) for severity, 1.40 (95% CI 1.20–1.64) for self-efficacy, and 1.79 (95% CI 1.45–2.22) for perceived benefits; all remained significant after Holm correction. These results support consistent, moderate, construct-specific correspondence between baseline real-life beliefs and subsequent in-game belief

### 3.4 Beliefs and Quarantine Behavior

Beliefs showed little robust association with observed quarantine behavior. In exploratory participant-level correlations, S1 severity (ρ=0.132, unadjusted p=0.0155) and S2 susceptibility (ρ=0.149, unadjusted p=0.0157) were positively associated with quarantine rate, but neither survived Holm correction across the eight belief–behavior comparisons. The remaining correlations were small and nonsignificant (**Figure 6**).

**Figure 6:**
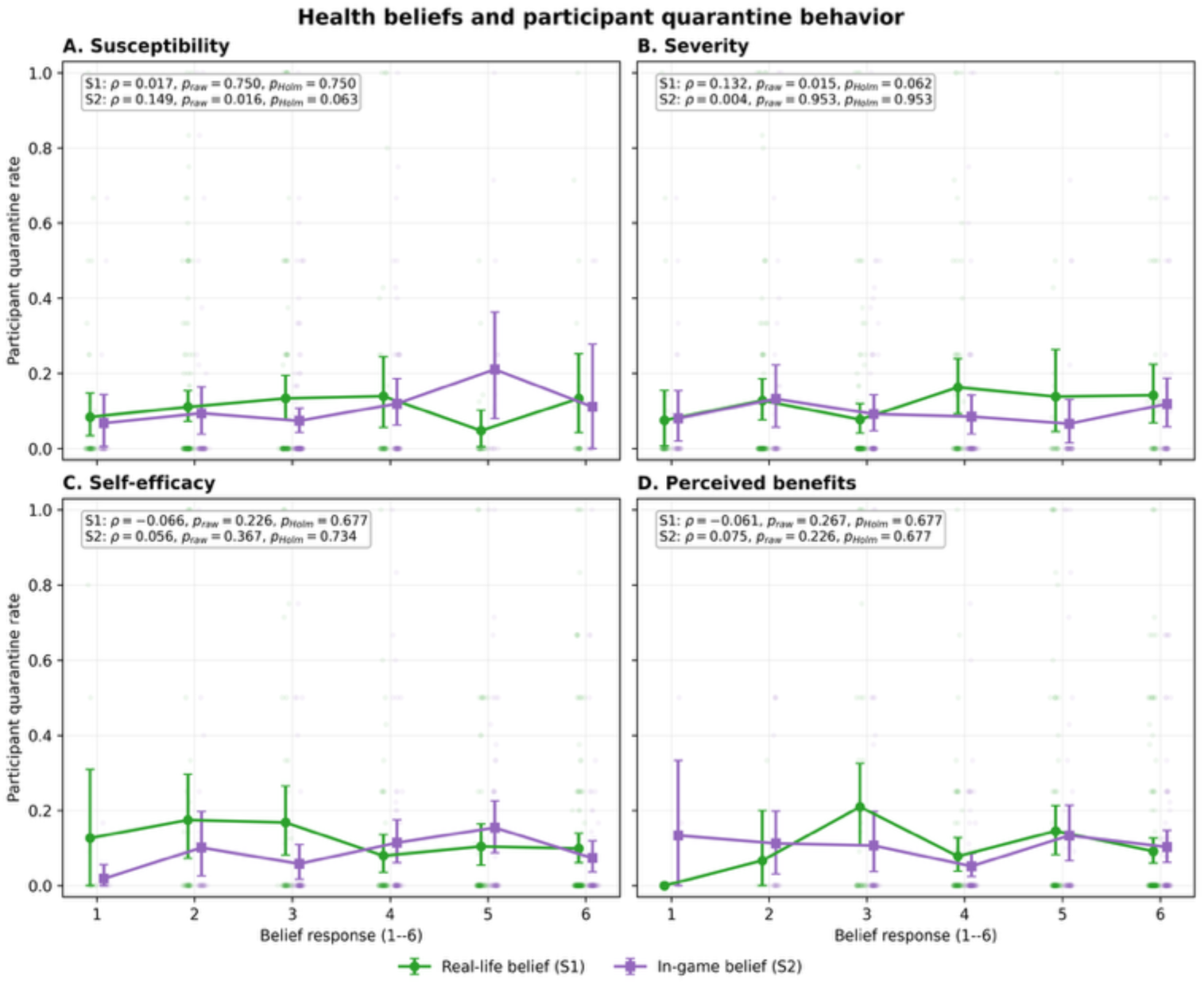
Beliefs and participant quarantine behavior. Unadjusted participant-level relationships between quarantine rate and susceptibility, severity, self-efficacy, and perceived benefits. Blue denotes S1 and red denotes S2. Insets report Spearman correlations with Holm-adjusted p-values.

In additive repeated-decision models, no belief item was consistently associated across the GEE and quasi-binomial specifications. The S1-adjusted GEE estimate for women was positive (p=0.046), but the corresponding quasi-binomial estimate was nonsignificant (p=0.147); S2-adjusted gender estimates were nonsignificant in both models.

### 3.5 Randomized Barrier Effect and Hypothesis Tests

Participants recorded 71 quarantine and 671 non-quarantine events in G1 (9.6%) and 78 quarantine and 795 non-quarantine events in G2 (8.9%). The randomized high-barrier condition did not detectably reduce quarantine in the overall behavioral cohort (**Figure 7A**). The full 15-day GEE estimated an OR of 0.90 for G2 versus G1 (95% CI 0.58–1.40, p=0.650). Excluding the four conflicting duplicate days gave a similar OR of 0.92 (p=0.718). The participant-level quasi-binomial estimate was also null (OR 0.93, 95% CI approximately 0.58–1.48, p=0.753), as were fractional-logit and OLS-HC3 sensitivity analyses. H1 was therefore not supported.

**Figure 7:**
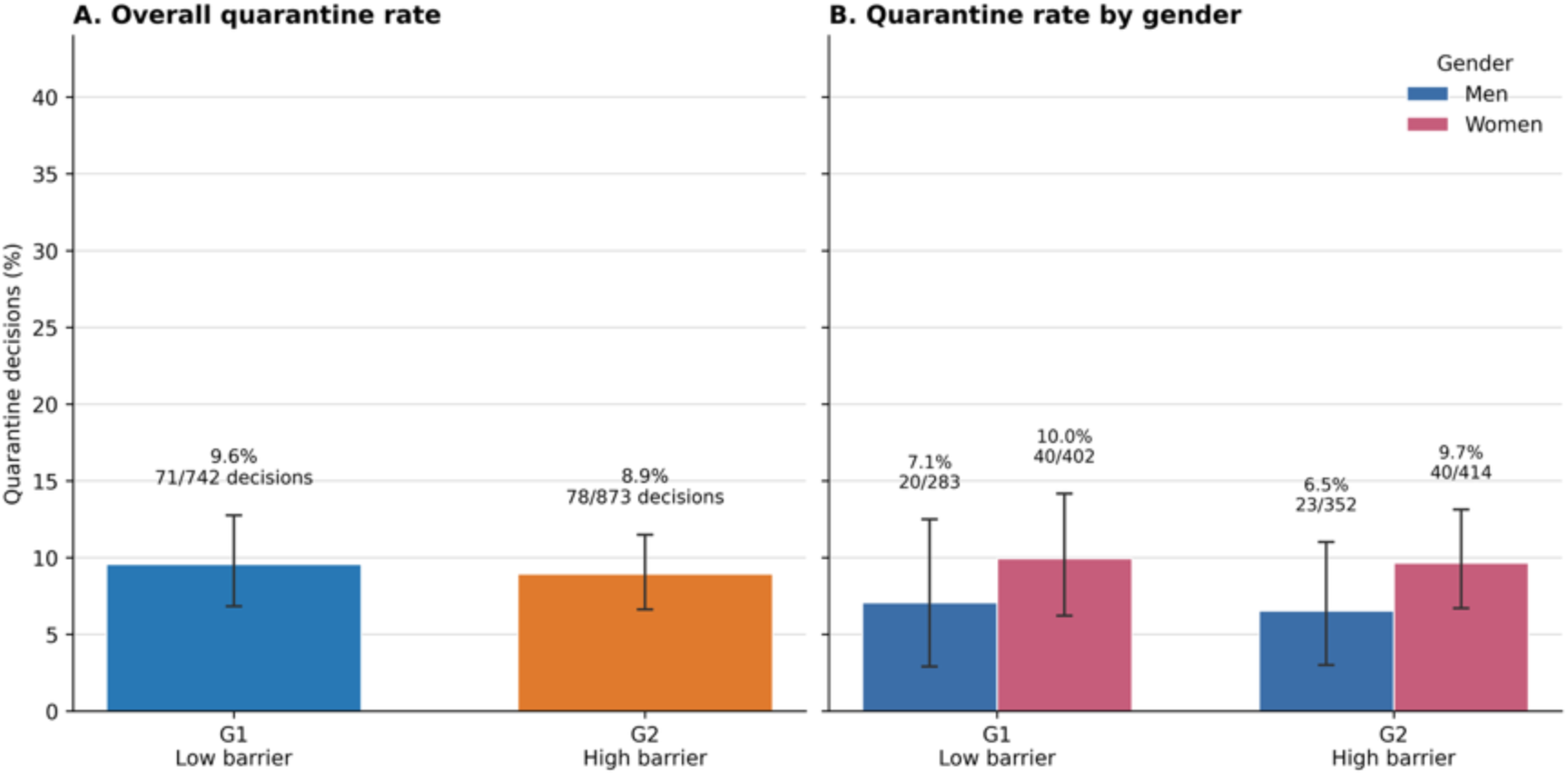
Observed quarantine-choice rates by randomized group. (A) Overall event-weighted rates. (B) Rates among men and women. Error bars are participant-cluster bootstrap CIs. These are descriptive; GEE and quasi-binomial models provide inference.

The ordinary Poisson and binomial variance assumptions underestimated the observed variation. Pearson scale estimates ranged from approximately 1.6 to 2.3 across the principal models. Among 663 consecutive-decision pairs, the probability of quarantine was 34.5% following a quarantine decision and 3.8% following an active decision (odds ratio 13.42, p<0.001). Exchangeable working correlations in the GEE models ranged from approximately 0.17 to 0.21. These diagnostics motivated inference from robust GEE and quasi-binomial standard errors rather than the original unscaled models.

The four baseline belief-by-group interactions were jointly null in both the GEE (Wald 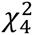 = 1.23, p=0.873) and quasi-binomial model (p=0.983), providing no support for H2. The S1 self-efficacy-by-group interaction was also null (GEE p=0.454; quasi-binomial p=0.561), providing no support for H3. Corresponding secondary models based on S2 beliefs likewise found no joint moderation (GEE p=0.212; quasi-binomial p=0.407) and no self-efficacy interaction (GEE p=0.832; quasi-binomial p=0.718).

A secondary contrast estimated the barrier effect for a participant scoring at the floor of all four belief scales. The S1 floor-profile contrast was null (GEE OR 0.71, p=0.734; quasi-binomial OR 0.72, p=0.774). For the S2 floor profile, the estimate was strongly negative but differed in inferential strength across specifications: GEE OR 0.060 (p=0.039) and quasi-binomial OR 0.058 (p=0.056). Because the joint S2 interaction test was null and the floor-profile result did not reach conventional significance under both final models, the earlier Poisson-based claim that the barrier selectively deterred skeptical participants was not considered robust.

### 3.6 Gender and Quarantine Adoption

Among participants with binary gender data, women had higher mean participant-level quarantine rates in both groups: 14.4% in G1 and 14.6% in G2, compared with 9.0% and 7.0% among men. Women were more likely than men ever to quarantine (unadjusted OR 2.72, p=0.0002; arm-adjusted OR 2.76, p=0.0002). The gender-by-group interaction for ever quarantining was null (OR 1.20, p=0.737), as were repeated-decision gender-by-group interactions. The evidence therefore supports a gender difference in adoption, not a differential response to the randomized barrier (**Figure 7B**).

### 3.7 Exploratory Exit-Survey Results

All S3 analyses involved 48 respondents contributing 402 participant-days. None of the four participant-level Spearman associations, group-adjusted GEE effects, quasi-binomial effects, or item-by-group interactions survived Holm correction. For S3-Q3 retrospective correspondence, the participant-level estimate was ρ=0.012 (Holm p=1.000); its GEE interaction OR was 1.65 (Holm p=0.482). These post-outcome analyses were underpowered, selected, and exploratory.

Nine additional participants submitted a separate voluntary feedback form. The median overall-experience rating was 3 of 5 (IQR 3–4), and the median ease-of-use rating was 6 of 10 (IQR 5–7). These nine submissions were not linked to trial outcomes and cannot estimate acceptability in the study population or support randomized-arm comparisons. Aggregate feedback is reported only as implementation context in the Supplementary Materials.

### 3.8 Exploratory Network Structure

The aggregate contact network included 442 participants with at least one retained edge and 3,295 edges. The observed weighted assortativity coefficients were largest for days not quarantined (r=0.46), S2 perceived benefits (r=0.48), S2 severity (r=0.40), and S1 susceptibility (r=0.23). These estimates describe similarity among connected participants but were not treated as confirmatory evidence: the exploratory resampling tests did not account for the full network dependence structure or multiplicity across attributes. Numeric assortativity for gender was not interpreted because its category codes do not form a continuous scale. The degree distribution was described graphically without claiming that the network was scale-free.

### 3.9 Summary of Findings

The trial established the operational feasibility of combining in-app randomization, repeated choices, Bluetooth contact measurement, and timed belief surveys in a naturalistic campus experiment. It supported matched correspondence between real-life and in-game beliefs but did not show that those beliefs reliably predicted quarantine choices. The randomized fixed barrier had no detectable average effect, and the preregistered joint moderation and self-efficacy hypotheses were unsupported. Baseline severity and the S2 floor-profile barrier contrast provided non-confirmatory signals for focused replication. Women were more likely to adopt quarantine at least once, without evidence of a gender-specific barrier effect. Exit-survey and voluntary-feedback findings remained exploratory.

## 4. Discussion

### 4.1 Principal Findings

The principal contribution of this study is methodological: it demonstrates that a preregistered randomized behavioral experiment can be embedded in an app-based gamified epidemic simulation while collecting longitudinal choices, passively sensed contacts, and prospectively timed belief measures. The trial recruited a large campus cohort, implemented randomized incentive conditions without researcher assignment, and linked experimental manipulation to repeated individual behavior in a naturalistic social environment. This establishes an empirical and technical foundation for iterative epigame RCTs, even though the present implementation did not confirm its behavioral hypotheses.

Substantively, the trial produced a clear distinction between transfer of stated beliefs and prediction of observed behavior. Participants’ baseline beliefs were consistently associated with the corresponding beliefs reported after entering the game. Yet individual beliefs explained little variation in quarantine, and neither baseline nor in-game beliefs jointly modified the fixed opportunity-cost effect.

The randomized high barrier also produced no detectable average change. Descriptive rates differed by less than one percentage point, and full-cohort GEE and quasi-binomial estimates agreed closely. A large S2 floor-profile estimate was significant only under GEE and was accompanied by null joint interaction tests. It is therefore a hypothesis-generating extreme-profile result, not evidence that the barrier selectively deterred skeptical participants.

The data nevertheless identify two signals worth carrying forward under explicit prospective hypotheses. Baseline severity had a small positive association with quarantine that narrowly missed significance after multiplicity correction. The S2 floor-profile contrast was large and directionally consistent with a gatekeeping mechanism in which a high opportunity cost particularly suppresses quarantine among participants with uniformly low in-game beliefs, but it was not robust across the two final model specifications. Neither pattern supports a claim of effect. Together with confirmatory signals in subsequent studies, however, they justify focused measurement, power calculations, and preregistered tests in subsequent implementations.

### 4.2 Belief Correspondence Without Established Behavioral Parallelism

The S1–S2 results provide evidence of construct-specific indicator correspondence. Relative differences among participants were preserved when the same four belief domains were reframed from respiratory infection and real-life quarantine to the virtual pathogen and in-game quarantine. Perceived benefits and self-efficacy showed the largest associations, while severity showed the weakest, but every matched relationship remained positive after Holm correction. The prospective ordering of S1 before S2 further supports interpretation as baseline beliefs predicting subsequent in-game reports, although it does not establish a causal effect.

This correspondence should not be equated with behavioral validity. The belief measures were associated with one another but generally did not predict what participants did when presented with daily quarantine choices. A person may carry a stable view about susceptibility or quarantine effectiveness into a game while making each choice in response to immediate points, infection status, social opportunities, accumulated experience, or other unmeasured considerations. The findings therefore distinguish correspondence of stated constructs from correspondence of behavior. The former was supported; the latter was not established by this study.

Measurement may also contribute to the disconnect. Each HBM domain was represented by one six-point item, limiting reliability and coverage of the underlying construct. Beliefs were measured at discrete survey times, whereas decisions occurred repeatedly and may have reflected changing information. Future studies could use validated multi-item scales and brief repeated measures collected near the decision window, with the timing and analysis specified prospectively.

### 4.3 Opportunity Cost and the Skeptic Profile

Increasing the point gap from 5 to 12 did not measurably change average quarantine propensity. This result concerns the implemented fixed-reward design and should not be interpreted as proving that opportunity costs are irrelevant. Participants may not have valued the added points as intended, may have communicated reward schedules across arms, or may have prioritized leaderboard and lottery outcomes differently. The low daily response density and technical variability may also have attenuated the randomized contrast.

The floor-profile result offers a more specific hypothesis for that work. If opportunity cost functions as a gatekeeper, its strongest effect should occur among participants who perceive little threat or benefit and therefore have less intrinsic motivation to quarantine, which is generally consistent with studies during COVID-19 that showed financial costs are a stronger predictor of self-isolation than intrinsic beliefs [40, 51]. Indeed, the current S2 contrast had the predicted direction and a large estimated magnitude, but borderline after the quasi-binomial correction. A subsequent trial should define this profile and contrast before data collection, ensure adequate observations across the low-belief region, and power the study for the interaction.

### 4.4 Gender and Quarantine Adoption

Women were more likely than men to quarantine at least once, a participant-level adoption result that was robust to adjustment for randomized arm and directionally consistent with prior evidence on gender differences in protective behavior [51]. Their mean quarantine rates were also higher in both arms. However, evidence that gender independently predicted repeated daily quarantine propensity depended on model specification, and no model supported a gender-by-barrier interaction. The findings are therefore more consistent with a gender difference in whether participants ever adopted quarantine than with a clearly estimated difference in the rate of repeated quarantine once opportunities and participant heterogeneity were considered. This distinction also cautions against treating the observed gender association as validation of real-world behavioral correspondence.

### 4.5 Network Findings

The contact network demonstrates the feasibility of combining passively sensed proximity data with randomized behavioral measurement. Several attributes had positive observed assortativity, including non-quarantine frequency and selected belief variables. These patterns are descriptive, however. The exploratory resampling approach did not preserve the complete dependence structure of the network, multiple attributes were examined, and numerical category codes are inappropriate for interpreting gender as a continuous assortative trait. The present analysis therefore does not establish social influence, homophily, or a scale-free topology. Confirmatory network analyses would require prespecified statistics and null models that preserve relevant network features.

### 4.6 Strengths and Limitations

Strengths include successful implementation of a preregistered app-based RCT, automated randomized incentives, repeated observed choices, prospectively measured beliefs, and naturalistic Bluetooth contact measurement. The design demonstrates that experimental behavioral hypotheses can be tested within an evolving, networked epidemic simulation rather than only through hypothetical survey responses. The use of full-cohort GEE and participant-level quasi-binomial models allowed uncertainty to reflect within-participant association and extra-binomial heterogeneity. The matched belief result was consistent across rank-based and ordinal methods.

Several limitations constrain interpretation. Only 403 of 545 participants with recoverable assignments made a choice. Conditioning on engagement may compromise randomization if prompt response depended on assignment and behavioral propensity. Participants contributed unequal numbers of decisions, and nearly three quarters never quarantined. Robust models improve variance estimation but do not remove informative nonresponse.

Bluetooth connectivity problems may have undercounted contacts and virtual transmission, while cross-arm communication may have weakened the intended contrast. The public leaderboard and lottery may also have emphasized point maximization over other game considerations.

These implementation limitations provide plausible explanations for attenuation, not demonstrated explanations for the null results. Bluetooth connectivity problems could reduce the salience or credibility of simulated exposure; unequal response to daily prompts reduced the effective behavioral sample and produced sparse longitudinal records; and a fixed reward schedule may have become predictable or been discussed across groups. Gameplay may also have been insufficiently varied or responsive to sustain attention over 15 days. The trial did not experimentally isolate these mechanisms or measure engagement continuously, so their effects cannot be separated from a genuinely small or absent treatment effect. They instead define concrete design targets for subsequent studies: more reliable proximity sensing, richer and more responsive gameplay, clearer feedback on consequences, stronger retention procedures, and prospective process measures of engagement and treatment fidelity.

Finally, regarding context parallelism: The quarantine mechanic as currently implemented—a digital button press—was designed to achieve indicator parallelism (a clear binary choice) while accounting for practical considerations (participants cannot be asked to physically quarantine) but because of this, it lacked the experiential weight of real-world isolation, thereby limiting context parallelism. To address this, future studies should focus on enhancing the salience of behavioral choices, ensuring that the decision “matters” psychologically to the player. This could be achieved by designing interactions that enforce “digital isolation” (e.g., temporary app lockout) or by fostering a deeper emotional attachment to the avatar. Such mechanisms would better mimic the non-monetary costs of quarantine (e.g., boredom, FOMO) and the emotional consequences of infection, thereby narrowing the gap between a gamified decision and a real-world dilemma.

## 5. Conclusion

The central contribution of this study is to establish a workable foundation for rigorous epigame RCTs. The platform combined experimental control with repeated behavior and measured social context at a scale that would be difficult to reproduce in a conventional laboratory. The study also shows why feasibility should not be conflated with validation: a platform may preserve meaningful variation in stated beliefs while observed choices remain governed by immediate game contingencies. Technical feasibility, survey correspondence, behavioral correspondence, and responsiveness to randomized mechanisms should therefore be evaluated separately.

At AUIB, real-life beliefs predicted their matched in-game counterparts, but neither those beliefs nor the fixed high barrier reliably predicted quarantine. The small severity association and large but specification-sensitive S2 floor-profile contrast provide disciplined hypotheses for future work, not confirmed evidence of a gatekeeper effect. Women were more likely to adopt quarantine, without evidence that the barrier operated differently by gender. Future multisite trials should improve technical reliability and sustained engagement while harmonizing outcome definitions, survey wording, incentive contrasts, attrition tracking, and robust model specifications before data collection. Claims about real-world protective behavior should remain contingent on direct behavioral evidence rather than inferred from similarity between survey responses.

## Data Availability

All data produced are available online at

https://doi.org/10.5281/zenodo.18209231

## Acknowledgments

The authors wish to thank the American University of Iraq – Baghdad for supporting this project. We would also like extend our gratitude to the AUIB students who volunteered to help during recruitment and logistics of the epigame: Waqas Saad Salih, Mostafa Nabeel Mohammed, Mohammedali Hussein Mihbas, Basmala Mazen Fakihir, Maryam Ahmed Naji, Daniah Ahmed Mawlood, Dur Ali Noaman, and Dimah Nayzak Jamal.

## Sources of funding

The project was supported with startup grant funds from AC, compute resources from the Broad Institute, and in-kind contributions from DK.

## Notes

### Competing Interest Statement

The authors have declared no competing interest.

### Clinical Trial

OSF Registration DOI 10.17605/OSF.IO/QEV6N

### Author Declarations

Ethical approval was granted by the Institutional Review Boards of the University of Massachusetts Chan Medical School and the American University of Iraq - Baghdad (study ID 2465)

### Summary of Updates

The methods are results were significantly updated by using (a) Holm-adjusted Spearman correlations, and prospective cumulative-logit models to asses belief correspondence, and (b) participant-day binomial generalized estimating equations (GEE) and participant-level quasi-binomial models to analyze in-game choices (which have temporal dependencies are they represent consecutive decisions made by the same individuals over the duration of the study). This reduced the statistical significance of several findings reported in the first version of the paper. Figures 3 through 7 are new to depict patterns in the data more clearly and reflect the updated results.

