## Supplementary materials for "App-based Epidemic Game to Model Belief-Behavior Mapping and Cost Incentives in Voluntary Quarantine: A Randomized Controlled Trial"

#### Evaluating Belief Correspondence and Fixed Cost Incentives in Voluntary Quarantine: A Randomized Controlled Trial Using an App-Based Epidemic Game at AUIB

Companion to the main AUIB manuscript draft.

**Source code:** https://github.com/colabobio/auib-epigame-analysis

#### Supplementary Methods

##### Analysis Populations and Outcome Construction

After removing registered participants who did not a group assignment (caused by a bug in the app that did not upload the group to the cloud server), we ended up with 545 participants randomized to either Group or Group 2. The behavioral population included 403 participants with at least one recorded quarantine choice: 188 in Group 1 (G1, low barrier) and 215 in Group 2 (G2, high barrier). Complete S1 belief data were available for 392 participants, complete S2 beliefs for 263, matched S1–S2 beliefs for 276, and complete S3 data for 48. Daily binary outcomes (quarantine and remain active) yielded 1,615 observations149 quarantine and 1,466 remain active.

##### Prospective S1-to-S2 Correspondence

For each domain, a cumulative-logit model estimated the odds of a higher in-game S2 response per one-point increase in its matched baseline S1 response, adjusting for randomized group. Expanded models included all four S1 domains and group. Responses retained their ordinal six-point scale. Holm correction was applied across the four matched-domain tests.

##### Final Models for the Randomized Barrier Hypotheses

Two complementary model families were used. Participant-day generalized estimating equations (GEE) used a binomial family, logit link, participant clustering, exchangeable working correlation, and robust standard errors. Participant-level quasi-binomial models used quarantine proportion as the outcome, total recorded choices as the binomial variance weight, and Pearson scaling (`scale='X2'`).

The arm-only effect used all 403 behavioral participants. S1 moderation models required complete S1 Questions 1–4 and binary gender and included 327 participants and 1,449 participant-days. S2 moderation models included 250 participants and 1,279 participant-days. In participant-level models, the corresponding complete-case denominators were 1,451 S1 choices and 1,280 S2 choices because the four conflicting duplicate days were retained as recorded events.

Beliefs retained their original Likert coding, with 1 as the reference value. Thus, the group coefficient in models containing belief-by-group interactions represents the group effect at the belief floor for men. Standardized floor-profile contrasts averaged the gender component over the relevant complete-case sample. H2 and its S2 extension were joint four-degree-of-freedom tests of belief-by-group interactions; H3 and its S2 extension tested self-efficacy-by-group interaction.

#### Exploratory Exit-Survey Analysis

The four S3 items were available for 48 participants contributing 402 participant-days:

- **S3-Q1, perceived realism:** higher values indicate greater perceived realism.
- **S3-Q2, concern:** higher values indicate greater concern about virtual infection.
- **S3-Q3, retrospective real-life correspondence:** higher values indicate greater reported similarity between game choices and intended real-life behavior.
- **S3-Q4, difficulty understanding information:** higher values indicate greater difficulty, not better understanding.

Participant-level Spearman correlations described unadjusted associations with quarantine rate. Group-adjusted associations were estimated with participant-day GEE and participant-level Pearson-scaled quasi-binomial models. Interaction models included the S3 item, group, and item-by-group interaction. Holm correction was applied separately across the four items for each declared family: Spearman associations, GEE main associations, quasi-binomial main associations, GEE interactions, and quasi-binomial interactions. These post-outcome analyses were not preregistered and cannot establish prospective prediction or causation.

#### Voluntary Post-Game Feedback

A separate feedback form received nine voluntary submissions. It was not linked to behavioral or randomized-group data. The analytic export excluded timestamps, direct identifiers, response-level rows, and verbatim comments. Overall experience used a five-point scale and ease of app use a ten-point scale.

Free-text responses were indexed using reproducible keyword rules within three overlapping domains: enjoyed elements, reported problems, and suggested improvements. A respondent could match multiple themes but was counted at most once per theme. Counts are primary; denominators are the numbers providing substantive text in the relevant domain. Only themes mentioned by at least three respondents are reported. This keyword indexing is descriptive and non-exhaustive and is not a validated qualitative analysis. There were only nine submissions, therefore results cannot estimate population prevalence or acceptability.

### Survey Instruments

#### Survey S1: Infectious Diseases in Real Life

**Description:** Choose the responses that best represent your beliefs. There is no right or wrong answer.

1. **Susceptibility:** How likely do you think it is that you will get infected with a respiratory virus (for example, influenza or COVID-19) in the next three months? 1 = virtually impossible; 6 = almost certain.
2. **Severity:** If you were infected with a respiratory virus in the next three months, how serious do you think the impact would be on your health? 1 = no impact at all; 6 = very significant impact.
3. **Quarantine self-efficacy:** How confident are you that you could successfully follow home quarantine during a new infectious disease with potentially serious health effects? 1 = not confident at all; 6 = completely confident.
4. **Perceived benefits:** How effective do you think home quarantine would be in preventing infectious disease spread? 1 = not effective at all; 6 = extremely effective.
5. **Gender:** male; female; other; prefer not to disclose.
6. **AUIB affiliation:** student; faculty; staff; other; prefer not to disclose.

#### Survey S2: Virtual Epidemic Game at AUIB

1. **Susceptibility:** How likely do you think it is that your avatar will become infected with the virtual pathogen? 1 = virtually impossible; 6 = almost certain.
2. **Severity:** If your avatar becomes infected, how much do you think this would affect your chance of winning a gift card in the point-based lottery? 1 = not affected at all; 6 = extremely affected.
3. **Quarantine self-efficacy:** How confident are you that you can successfully follow in-game quarantine? 1 = not confident at all; 6 = completely confident.
4. **Perceived benefits:** How effective do you think in-game quarantine is in preventing spread of the virtual pathogen? 1 = not effective at all; 6 = extremely effective.

#### Survey S3: Experience During the AUIB Epidemic Game

1. How well do you think the game represents infectious disease transmission in real life? 1 = very poorly; 6 = extremely accurately.
2. Were you worried about becoming virtually infected during the game? 1 = not worried at all; 6 = very worried.
3. How closely did your quarantine decisions during the game reflect what you would do in a real-life outbreak? 1 = not at all similar; 6 = exactly what I would do.
4. How difficult was it to understand the information received during the game? 1 = very easy to understand; 6 = extremely hard to understand.

5. Field of study: healthcare or biology; law, international studies, or English literature; education or psychology; business; engineering or chemistry; computer science.

#### Supplementary Results

##### Supplementary Table S1. Matched S1-to-S2 Belief Correspondence

| Domain | Matched $N$ | Spearman $\rho$ | Holm $p$ | Prospective cumulative-logit OR (95% CI) | Interpretation |
|---|---|---|---|---|---|
| Susceptibility | 257 | 0.204 | 0.0020 | 1.32 (1.11–1.57) | Higher baseline susceptibility predicted a higher in-game response |
| Severity | 257 | 0.126 | 0.0431 | 1.21 (1.02–1.44) | Small positive matched-domain association |
| Self-efficacy | 257 | 0.267 | $< 0.001$ | 1.40 (1.20–1.64) | Positive matched-domain association |
| Perceived benefits | 257 | 0.353 | $< 0.001$ | 1.79 (1.45–2.22) | Largest matched-domain association |

*Notes:* ORs represent the cumulative odds of a higher S2 response per one-point increase in the matched S1 item, adjusting for randomized group. The Spearman and prospective ordinal results each survived Holm correction across four domains. Positive association does not imply equality of S1 and S2 responses or behavioral validity.

##### Supplementary Table S2. Final Tests of the Randomized Barrier Hypotheses

| Hypothesis or estimand | Analysis sample | GEE result | Quasi-binomial result | Decision |
|---|---|---|---|---|
| Overall G2 versus G1 effect | 403 participants; 1,611 participant-days / 1,615 choices | OR 0.90 (0.58–1.40), $p = 0.650$ | OR 0.93 (0.58–1.48), $p = 0.753$ | No detectable average effect |
| H2: joint S1 belief-by-group interactions | 327 participants; 1,449 participant-days / 1,451 choices | Wald $\chi^2_4 = 1.23$, $p = 0.873$ | Wald $\chi^2_4 = 0.39$, $p = 0.983$ | Not supported |

| Hypothesis or estimand | Analysis sample | GEE result | Quasi-binomial result | Decision |
|---|---|---|---|---|
| H3: S1 self-efficacy-by-group interaction | Same S1 complete-case sample | $p = 0.454$ | $p = 0.561$ | Not supported |
| S1 floor-belief profile | Same S1 complete-case sample | OR 0.71, $p = 0.734$ | OR 0.72, $p = 0.774$ | Not supported |
| Joint S2 belief-by-group interactions | 250 participants; 1,279 participant-days / 1,280 choices | Wald $\chi^2_4 = 5.84$, $p = 0.212$ | Wald $\chi^2_4 = 3.99$, $p = 0.407$ | Not supported |
| S2 self-efficacy-by-group interaction | Same S2 complete-case sample | $p = 0.832$ | $p = 0.718$ | Not supported |
| S2 floor-belief profile | Same S2 complete-case sample | OR 0.060, $p = 0.039$ | OR 0.058, $p = 0.056$ | Specification-fragile; not confirmed |

*Notes:* G1 is the low-barrier reference group. The overall randomized comparison is distinct from the floor-profile contrasts. The floor profile assigns a value of 1 to all four belief items and averages over the sample gender distribution. Null joint S2 interaction tests and disagreement at the conventional threshold make the S2 floor result inconclusive.

#### Supplementary Table S3. Marginal Belief Associations with Participant Quarantine Rate

| Survey and domain | $N$ | Spearman $\rho$ | Raw $p$ | Holm $p$ |
|---|---|---|---|---|
| S1 susceptibility | 336 | 0.017 | 0.7502 | 0.7502 |
| S1 severity | 336 | 0.132 | 0.0155 | 0.0619 |
| S1 self-efficacy | 336 | -0.066 | 0.2258 | 0.6773 |
| S1 perceived benefits | 336 | -0.061 | 0.2669 | 0.6773 |
| S2 susceptibility | 263 | 0.149 | 0.0157 | 0.0627 |
| S2 severity | 263 | 0.004 | 0.9525 | 0.9525 |
| S2 self-efficacy | 263 | 0.056 | 0.3669 | 0.7337 |
| S2 perceived benefits | 263 | 0.075 | 0.2256 | 0.6769 |

*Notes:* Holm adjustment was applied within S1 and S2. No association survived correction. These participant-level comparisons are exploratory and unadjusted.

#### Supplementary Table S4. Gender and Quarantine Adoption

| Estimand | Result |
|---|---|
| Women versus men, ever quarantined, unadjusted | OR 2.72, $p = 0.0002$ |
| Women versus men, ever quarantined, adjusted for group | OR 2.76, $p = 0.0002$ |
| Women versus men in adoption interaction model | OR 2.50, $p = 0.0231$ |
| Gender-by-group interaction for adoption | OR 1.20, $p = 0.7369$ |
| S1 additive GEE gender effect | $\beta = 0.540$ (95% CI 0.009–1.072), $p = 0.046$ |
| S1 additive quasi-binomial gender effect | $\beta = 0.426$ (95% CI -0.149–1.001), $p = 0.147$ |
| S2 additive GEE gender effect | $\beta = 0.492$ (95% CI -0.203–1.186), $p = 0.165$ |
| S2 additive quasi-binomial gender effect | $\beta = 0.377$ (95% CI -0.306–1.060), $p = 0.279$ |

*Notes:* Gender was coded as 0 for men and 1 for women. Participants with another or unavailable response were excluded only from analyses requiring the binary variable. The gender-by-group interactions in repeated-decision models were also null. The most consistent result concerns adoption at least once, not repeated daily propensity.

#### Supplementary Table S5. Exploratory S3 Associations with Quarantine

| Item | $N$ | Spearman $\rho$; raw / Holm $p$ | GEE group-adjusted OR (95% CI); raw / Holm $p$ | Quasi-binomial OR (95% CI); raw / Holm $p$ | GEE item-by-group OR; raw / Holm $p$ |
|---|---|---|---|---|---|
| S3-Q1 realism | 48 | 0.153; 0.300 / 0.901 | 1.20 (0.93–1.55); 0.166 / 0.665 | 1.20 (0.79–1.83); 0.399 / 1.000 | 1.11; 0.694 / 1.000 |
| S3-Q2 concern | 48 | 0.019; 0.896 / 1.000 | 0.92 (0.71–1.19); 0.524 / 1.000 | 0.92 (0.68–1.24); 0.586 / 1.000 | 0.87; 0.619 / 1.000 |
| S3-Q3 retrospective correspondence | 48 | 0.012; 0.936 / 1.000 | 0.93 (0.68–1.28); 0.672 / 1.000 | 0.94 (0.65–1.36); 0.732 / 1.000 | 1.65; 0.120 / 0.482 |
| S3-Q4 difficulty understanding | 48 | 0.224; 0.126 / 0.503 | 1.22 (0.80–1.86); 0.361 / 1.000 | 1.23 (0.83–1.82); 0.311 / 0.892 | 0.64; 0.297 / 0.892 |

*Notes:* Regression ORs represent a one-point increase. GEE analyses include 402 participant-days. No result survived Holm correction. S3 was collected after gameplay from a selected subset.

#### Supplementary Table S6. Aggregate Voluntary Feedback Ratings and Reportable Themes

| Domain | Rating or indexed theme | Aggregate result |
|---|---|---:|
| Overall experience | Five-point rating | $N = 9$; mean 3.11 (SD 1.05); median 3 (IQR 3–4); range 1–4; 4/9 rated 4 or 5 |
| Ease of app use | Ten-point rating | $N = 9$; mean 6.00 (SD 1.22); median 6 (IQR 5–7); range 4–8; 1/9 rated 8–10 |
| Enjoyed elements | Points and rewards | 4/7 substantive responses |
| Reported problems | Bluetooth or contact detection | 4/8 substantive respondents |
| Reported problems | Performance, loading, or crashes | 3/8 substantive respondents |
| Suggested improvements | Avatar or customization | 5/8 substantive respondents |
| Suggested improvements | Rewards, shop, or items | 5/8 substantive respondents |
| Suggested improvements | Realism, information, or feedback | 3/8 substantive respondents |

*Notes:* The form received nine voluntary submissions. Theme counts overlap and describe keyword-indexed mentions, not prevalence, sentiment intensity, or qualitative importance. Only themes with at least three mentions are shown. Counts are primary; percentages are omitted because the denominators are very small. No response-level or verbatim data are reported.

### Supplementary Figures

#### Supplementary Figure S1. Willingness-to-Accept Pilot

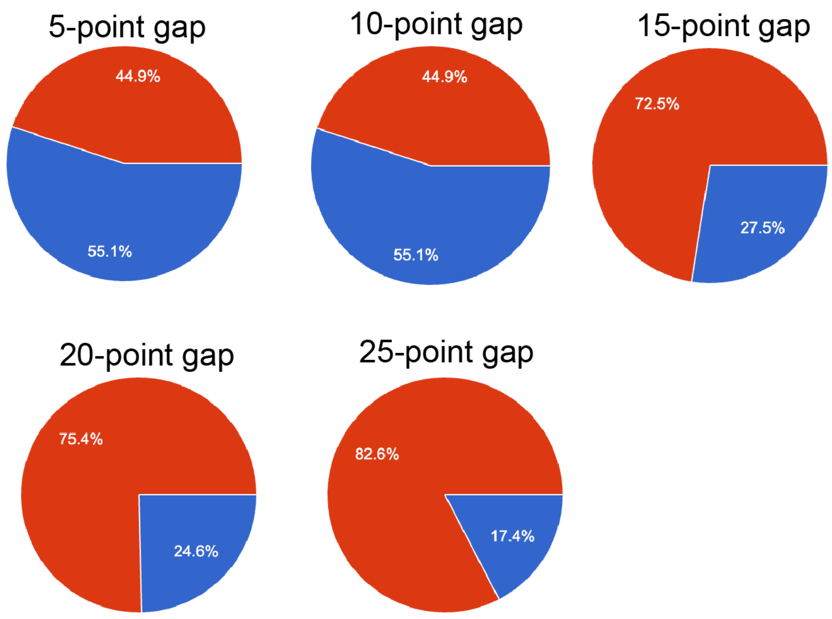


*Supplementary Figure S1. Willingness-to-Accept Pilot.*

**Supplementary Figure S1. Willingness-to-Accept Pilot.** Each pie chart represents the fraction of participants who picked the no quarantine option (red) versus quarantine (blue) for the given point gap.

#### Supplementary Figure S2. Voluntary Post-Game Ratings

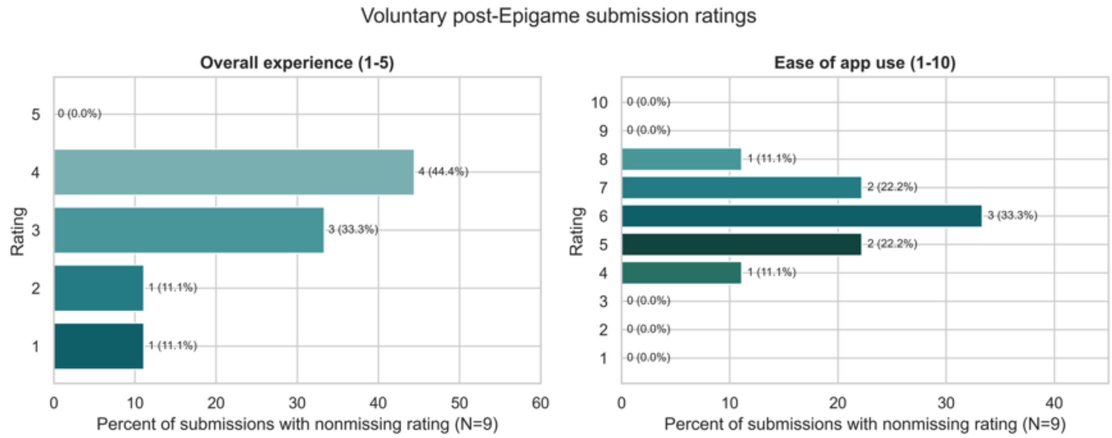


*Supplementary Figure S2. Voluntary post-game ratings.*

**Supplementary Figure S2. Voluntary post-game ratings.** Distribution of overall-experience ratings on a five-point scale and ease-of-use ratings on a ten-point scale across nine voluntary submissions. Bars retain each observed response category. The figure describes respondents only and is not an estimate of study-population acceptability.

#### Supplementary Figure S3. Network Analysis

Panel A shows the aggregate contact network (where weighted edges were calculated by adding up the total contact time between each pair of participants). Panel B shows the degree distribution (left is histogram of the node degree, right is the probability plot in log-log scale). Panel C shows the network colored by significantly assortative variables.

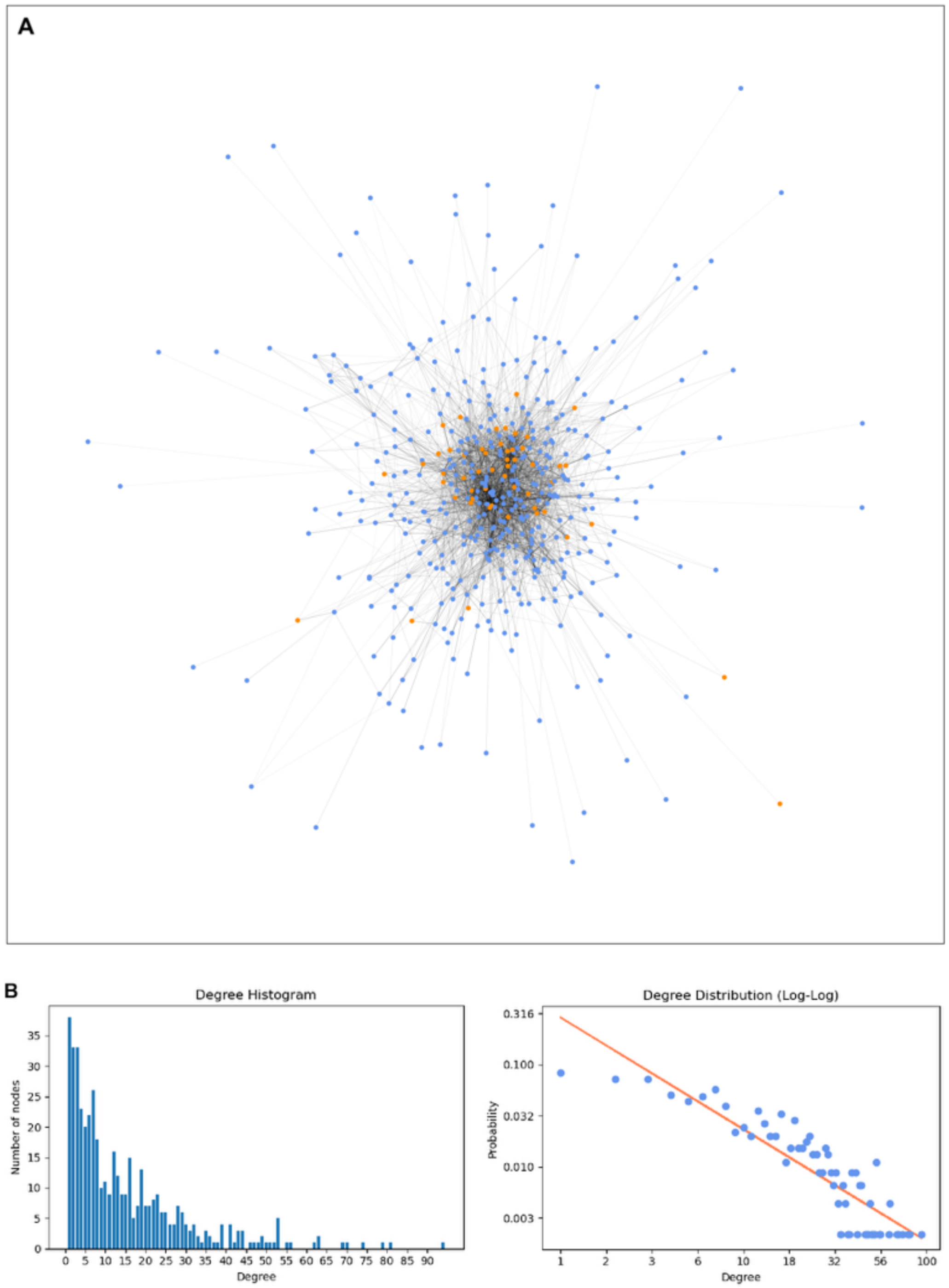

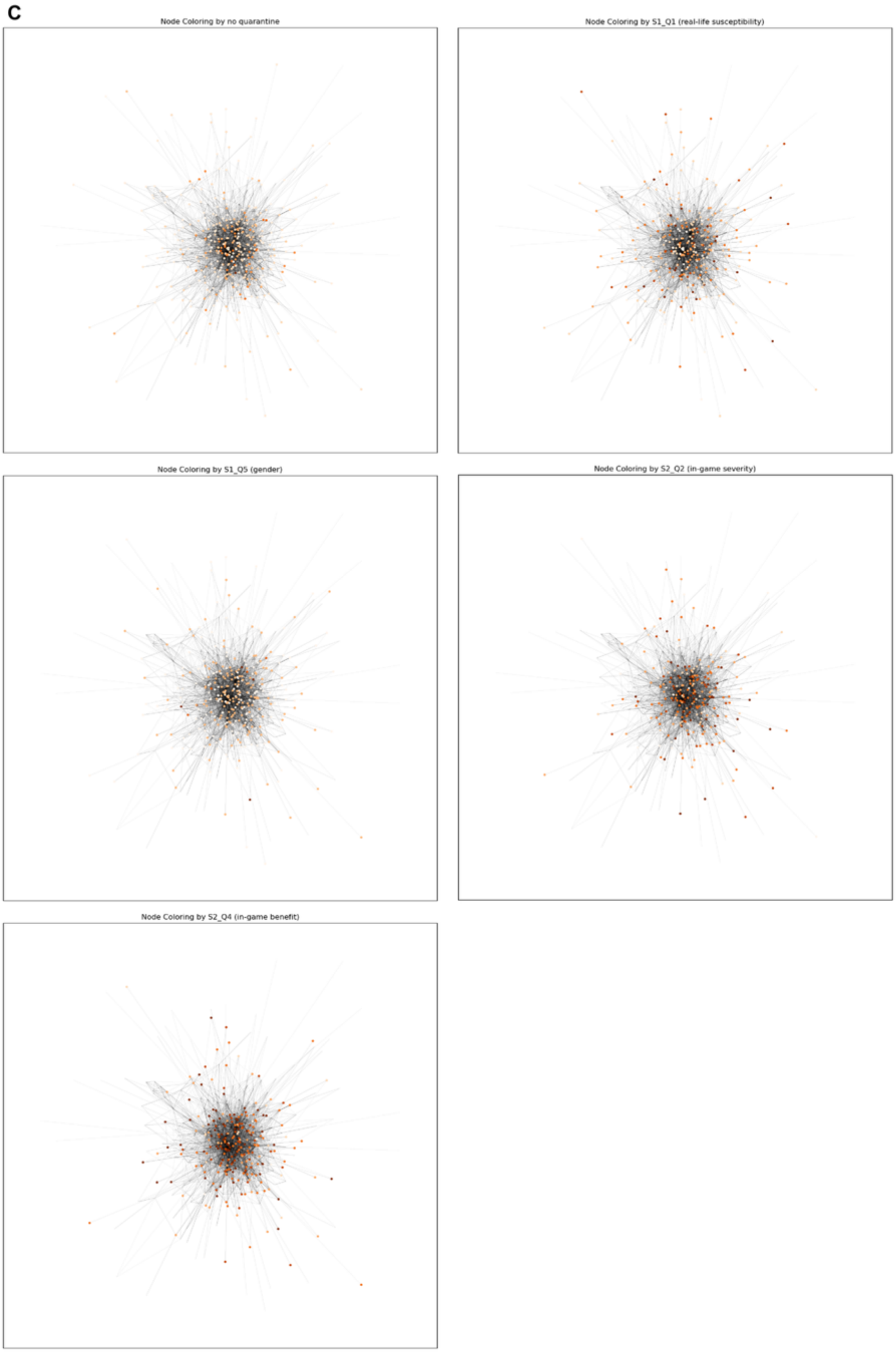
C
Node Coloring by no quarantine
Node Coloring by S1_Q1 (real-life susceptibility)
Node Coloring by S1_Q5 (gender)
Node Coloring by S2_Q2 (in-game severity)
Node Coloring by S2_Q4 (in-game benefit)
